# Development and clinical application of a consonant confusion task to evaluate hearing aid benefit

**DOI:** 10.64898/2026.04.23.26351598

**Authors:** Joshua J. Hajicek, Sara E. Harris, Stephen T. Neely

## Abstract

**Purpose:** This research sought to develop a low-cognitive-load speech-in-noise test based on consonant confusions with the potential for assessing hearing-aid benefit.

**Methods:** Vowel-consonant-vowel (VCV) stimuli with added speech-shaped noise were presented as a closed-set consonant identification task. Initially, consonant-confusion matrices were used to select, from a larger set of consonants and vowel contexts, a set of ten consonants and associated signal-to-noise ratios (SNR) that were sensitive to hearing loss. The sensitivity of the qVCV test to hearing loss was validated by comparing predicted pure-tone average (PTA) hearing thresholds with their audiometric PTA. Clinical viability of the qVCV test was assessed by comparisons to the QuickSIN test. Hearing-aid benefit was assessed by comparing test scores in unaided and aided conditions.

**Results:** The consonants most sensitive to hearing loss were /b d g t k v z s ʃ n/ in the vowel context /ɑ/. A cross-validated prediction of PTA had a mean-absolute error of 5.7 dB. The repeatability of qVCV at 50 trials was equivalent to the QuickSIN average of two lists. Hearing-aid benefit was quantified as a decibel reduction in hearing loss.

**Conclusions:** qVCV and QuickSIN performed similarly when test times are equated. The advantages of qVCV include lower cognitive demand, fewer learning effects, and automated scoring. PTA predicted by qVCV which greatly exceeds audiometric PTA may indicate either cognitive deficits or cochlear neural degeneration. The qVCV quantification of hearing-aid benefit may have clinical value.

## Introduction

Many individuals with self-reported “perceived” hearing difficulties suffer similar disruptions to daily life compared to individuals with clinically diagnosed hearing loss. Untreated hearing loss is estimated to increases the risk of dementia by up to 42%, but this increased risk may be mitigated by using hearing aids (Amieva et al., 2018; Marinelli et al., 2022). Standard pure-tone audiometry primarily measures threshold sensitivity and often fails to capture suprathreshold deficits, such as cochlear synaptopathy or temporal processing deficits, which contribute to speech-in-noise difficulties (Beck & Danhauer, 2019). Consequently, patients with ‘normal’ or near-normal thresholds but significant perceived difficulty may be overlooked. These limitations in the current standard of care indicate the need for improved, efficient hearing tests that can identify deficits beyond the audiogram.

Speech-in-noise (SIN) testing provides relevant insights into hearing function (Davidson et al., 2021). According to the American Speech-Language-Hearing Association (ASHA) and the American Academy of Audiology (AAA), audiologists should administer speech-in-noise tests as part of their standard test battery, but only 15% of practicing audiologists perform these tests (Beck & Danhauer, 2019). This raises concerns about how many patients missed by conventional audiometry have unexplained hearing difficulties and are not offered appropriate interventions at the time of their first visit.

Multiple standardized SIN tests exist, each differing in stimuli, language dependence, scoring, and clinical purpose. For example, the HINT (Hearing in Noise Test; (Nilsson et al., 1994)) is an adaptive test that uses sentences in speech-shaped noise and has been shown to be useful for directional microphone testing. The WIN (Words in Noise test; (Wilson et al., 2007)) is a simple task with monosyllabic words in babble, helpful when wanting to remove linguistic redundancy. The DIN (Digits in Noise test; (Schimmel et al., 2024)) is language-independent and uses digit triplets in noise. It is now available in app format and is useful for screening, especially in remote settings. One of the most widely adopted SIN tests in the clinical setting is the QuickSIN test (Killion et al., 2004). As implied in the name, test administration is “quick”, and it can be used as a functional assessment of SIN performance with hearing aids. Each SIN test likewise presents specific limitations, including concerns about administration time, variability across individuals, cognitive requirements, and the scoring and interpretation of outcomes.

Speech recognition tasks assess the combined auditory and linguistic processing abilities as well as cognitive functions. Recognition of speech in noise is difficult and requires multiple physical and neural processes that can begin to weaken or are lost as hearing decreases and age increases, causing speech clarity to decrease. Cognitive functions, such as working memory, are likely to account for much of the variability in many speech-in-noise tests (e.g., Akeroyd, 2008; Shinn-Cunningham & Best, 2008). The efficiency of a speech-in-noise test may be reduced by dependence on working memory and cognitive load when these abilities are impaired.

QuickSIN is composed of IEEE sentences that are presented in multi-talker babble noise (“IEEE Recommended Practice for Speech Quality Measurements,” 1969). In each list of six sentences, the signal-to-noise ratio (SNR) starts high and then decreases with each successive sentence, making it more difficult to understand and identify the target words. The final score is converted to an SNR loss score and based on this score, the degree of SNR loss is quantified (L Beck & L Danhauer, 2019). In the QuickSIN manual, SNR loss is defined “as the dB increase in the signal-to-noise ratio required by a hearing-impaired person to understand speech in noise, compared to someone with normal hearing (*QuickSIN Speech-in-Noise Test User Manual*, 2001).” SNR loss is used to evaluate how much the patient is struggling to hear in background noise and to help drive counseling the patient on realistic expectations when being fitted with hearing aids is recommended.

QuickSIN sentences were designed to be phonemically balanced; however, some studies have raised doubts about this. In a study by McArdle and Wilson (2006), the homogeneity of all 18 lists on the QuickSIN were assessed and showed that there was high variability in recognition performance among the lists, meaning that the results were not consistent with each other. This inconsistency between lists makes it difficult to quantify SNR loss and its prediction of hearing difficulties and hearing aid outcomes. Another factor to consider is the context eeect (meaning, grammar, prosody, etc.). QuickSIN may not accurately test speech perception because patients can recognize semantic and phonetic cues to guess what a word is, leading to inflated SNR scores (Phatak et al., 2009). Additionally, QuickSIN requires active recall and short-term memory to repeat sentences out loud. The QuickSIN relies on working memory because it requires remembering multiple words while ignoring background noise (Larsby et al., 2008). This can cause greater fatigue during testing which can decrease score reliability.

In addition to SIN tests, questionnaires are useful tools for identifying the self-perceived hearing difficulties experienced in daily life outside of the clinic. The Speech, Spatial and Qualities of Hearing Scale (SSQ) and it’s short form, SSQ-12, is commonly used in clinical settings and is strongly correlated with pure-tone hearing thresholds and measures of speech perception in noise (Motlagh Zadeh et al., 2025). Compared to other surveys of hearing disability in adults, such as the Abbreviated Profile of Hearing Aid Benefit (APHAB, (Cox & Alexander, 1995)) or the Hearing Handicap Inventory for Adults ( HHIA, (Newman et al., 1990)), the SSQ-12 is appropriate for use on people with a range of hearing thresholds, including normal hearing, because it does not ask questions regarding audibility and it has been used in prior research to study self-reported hearing in people with normal audiometric thresholds (Humes et al., 2013; Toscano & Allen, 2014).

Our primary motivation was to develop a clinically efficient tool to 1) predict PTA to identify “excess” hearing loss (deficits beyond what is predicted by the audiogram) and 2) quantify hearing aid benefit using a low-cognitive-load task. A consonant confusion task is well-suited for evaluating speech-perception abilities because consonant identification relies heavily on fine-grained acoustic cues—such as temporal precision, spectral detail, and rapid onset transitions—that are particularly vulnerable to suprathreshold auditory deficits. These cues are more sensitive to subtle peripheral or central auditory impairments than vowel-based or sentence-level materials. As a result, consonant-confusion stimuli are an effective and sensitive tool for detecting deficits that may not be captured by traditional audiometric or sentence-in-noise measures. The development of the “quick consonant-confusion test” (qVCV), employing noise-masked consonants in a single-vowel context and single-item closed-set trials to reduce variability and cognitive load, is detailed below.

A key translational benefit of the qVCV test is its ability to quantify hearing deficits that are greater than expected from the predicted pure-tone average (PTA), providing information not evident from the audiogram alone. Unlike other commonly employed SIN tests, the qVCV testing approach was designed to reduce reliance on working memory and cognitive load. Considering these advantages, another valuable application of the qVCV test may be to evaluate hearing aid benefit. Because the predicted PTA obtained in an unaided condition can identify greater hearing deficits than expected based on the audiogram alone, it may also support a recommendation for amplification intervention not otherwise indicated. The qVCV test can also be administered in aided conditions where the aided benefit can be quantified and the benefit between multiple devices, prescriptions, signal processing strategies, and settings could be compared which may lead to improved clinical outcomes and patient satisfaction.

The development of the qVCV test spanned across two experimental phases. The goal of Phase 1 was to demonstrate the ability to measure consonant confusion matrices in normal-hearing listeners and estimate SNRs that normalize performance across VCVs to achieve sensitivity to hearing loss. Phase 2 aimed to evaluate the clinical viability of qVCV with assessments of test repeatability and efficiency compared to the QuickSIN test of speech-in-noise perception, and to explore potential clinical applications of the qVCV test including evaluation of hearing-aid benefit.

## Methods

### Experimental Overview

Four independent experiments with different participants contributed to the development of the qVCV test. Phase 1 included Experiments 1 and 2; Phase 2 included Experiments 3 and 4.

Experiment 1 demonstrated the feasibility of measuring consonant-confusion matrices in normal-hearing listeners (n=20) and determined SNRs for each VCV that normalized performance to 90% for normal hearing listeners. The initial stimulus set included 234 VCV tokens (13 consonants × 3 vowels × 2 recordings × 3 SNRs). In Experiment 1, the SNRs were fixed at 0, 6, and 12 dB. SNR90 was extrapolated for each VCV and a narrower range, ±3 dB SNR, which were likely to encompass the SNR90 were selected for Experiment 2. The vowel context /u/ produced the highest variability in Experiment 1. Furthermore, performance on /u/ was largely collinear with /i/, offering limited additional diagnostic value. Therefore, /u/ was removed from further consideration to mitigate redundancy and reduce variability.”

In Experiment 2, consonant confusion matrices were acquired for 13 consonants in 2 vowel contexts, /ɑ i/, from 60 participants with normal hearing (n=15) and a range of hearing loss (n=45). Using logistic regressions, the VCVs that best predicted hearing status as normal or impaired were selected. The stimulus set was further reduced by examining the AUC for the separation of ears with normal hearing from ears with hearing loss, which reduced the number of tokens from 156 (13 consonants x 2 vowels x 2 recordings x 3 SNRs) to 20 (1 vowel x 10 consonants x 2 recordings x 1 SNR). The final set of 10 VCVs were, by design, optimally sensitive to hearing loss.

Experiment 3 validated the 10-consonant qVCV test procedures in a new group of 20 participants with normal hearing (n=10) and hearing loss (n=10),and in an aided condition to determine its ability to measure hearing aid benefit in the participants with hearing loss.

In a new cohort of 40 participants grouped by degree of hearing loss (normal (n=10), slight (n=10), mild (n=10), moderate (n=10)), Experiment 4 addressed two specific aspects: (1) assessment of test repeatability and efficiency compared to the QuickSIN test and (2) quantification of hearing aid benefit with lab-fit (n=20) and personal (n=9) hearing aids.

### Stimuli

Speech waveforms from a previous study (Nishi et al, 2017) were modified for this study. They included recordings of two productions each of 39 VCV tokens, consisting of 13 consonants /b d ɡ p t k v z f s ʃ m n/ symmetrically framed by one of three vowels /i ɑ u/ (e.g., /ɑkɑ/, /iki/, /uku/). To minimize variance due to age-related lexical access and across-linguistic backgrounds, the vowel contexts /θ ð ʧ ʤ l ɹ/ were not considered (Goldstein et al., 2005; Nishi et al., 2017; Sander, 1972). Each of the 78 VCVs (13 consonants × 3 vowels × 2 recordings) was spoken by an adult female talker in American English with a Midwestern accent. Each production was recorded in quiet and similar stress was used for the first and final vowels across the tokens (Nishi et al., 2017). The level of each VCV token was always fixed at 65 dB SPL (by setting its waveform rms value equal that of a 1-kHz tone at 65 dB SPL). VCV tokens were preceded by 25 milliseconds of silence and played until the final vowel decayed naturally into the noise floor of the recording. The increased difficulty associated with this sound level in individuals with hearing loss was intentional.

Each VCV token was mixed with long-term average speech-spectrum noise (Stelmachowicz et al., 1993). The frequency spectrum of the speech-shaped noise was equivalent to the long-term average of the talker but with a randomized phase. A detailed description of the long-term average speech-shaped noise can be found in McCreery et al. (2014). The level of the speech-shaped noise was adjusted for each VCV token to achieve the signal-to-noise ratios (SNRs) designated in each experiment.

### Equipment

The participants were seated in a sound-treated double-walled audiometric booth (IAC Acoustics, Naperville) at a desk with a PC monitor. A custom graphical user interface (GUI) software written in MATLAB (MathWorks, Natick, MA, USA) was used to present stimuli and record responses. A RME BabyFace Pro USB audio interface connected to a Microsoft Windows PC was used to control playback. The VCV stimuli were presented monaurally via an ER3A insert phone in Phase 1 (Etymotic Research, Elk Grove, IL) or Sennheiser HD 280 Pro circumaural headphones (Sennheiser electronic, Wedemark, Germany) in Phase 2. The output level was calibrated by measuring a 1000 Hz pure tone with a sound level meter (Larsen Davis SoundTrack LxT1) and GRAS artificial ear (Type 43AC occluded-ear simulator ear for insert earphones in Phase 1 Experiments and Type 43AA for circumaural headphones in Phase 2 Experiments) prior to data collection, and the voltage of the calibration tone was checked daily using a multimeter.

### Procedure

Following the presentation of each VCV token, the participants were instructed to choose the consonant that best matched what they heard. Choices consisted of a closed set of 14 buttons labeled with the consonants “b, d, ɡ, p, t, k, v, z, f, s, sh, m, n” and “other” (Phase 1) or “???” (Phase 2), which was displayed on a PC monitor. The labels of the buttons did not change with vowel context. Participants were instructed to only select “other” or “???” if they heard a consonant that was not a choice. Responses were scored as correct or incorrect. Selection of ‘other’ or ‘???’ was scored as an incorrect response. Performance was calculated in percentage correct, which was then converted to a Predicted PTA (PPTA). PPTA can be used to calculate residual hearing loss (RHL), which is defined as the difference between audiometric and predicted PTA.

Before the experiment, the participants were familiarized with the test in a practice block that consisted of a single presentation of each VCV token in quiet. Feedback was provided to indicate whether the response was correct or incorrect. After the practice block, the tester answered any questions and re-instructed if needed. During the experiment, stimuli were presented in test blocks where VCV tokens were mixed with noise, and no feedback was given. Tokens were presented only once and could not be repeated. The test was not timed; the next token was played after each response, and a progress bar was displayed on the screen.

### Participants

Participants were recruited through a database of volunteers who agreed to be contacted about research maintained by the Boys Town National Research Hospital (BTNRH). Participants were required to have normal hearing or mild-to-moderate sensorineural hearing loss and normal middle-ear status. Exclusion criteria included any known or diagnosed cognitive or neurological disorders and non-native speakers of English. The BTNRH Institutional Review Board approved all research protocols before recruitment, consent, screening, and data collection. The participants were compensated for their time.

All participants were asked three yes/no questions about their history of noise exposure, tinnitus, and hyperacusis via interview. In Phase 2, six questions from the SSQ-12 questionnaire were selected for their speech-in-noise context and used to quantify hearing difficulties in everyday listening situations. The modified SSQ questionnaire was administered via REDCap (Harris, PA et al.) on a tablet. If the participants were hearing aid users, they were instructed to answer as if they were not wearing hearing aids.

Outer-ear status was assessed by otoscopy to ensure that the ear canals were clear of obstructions, tympanic membranes were visually healthy, and the presence of any outer ear abnormalities could be ruled out. Middle-ear function was assessed with standard clinical tympanometry at 226 Hz using a Madsen Zodiac tympanometer. All participants had normal middle-ear status determined by air-bone gaps (ABG) ≤ 10 dB HL from 0.5 to 4 kHz and a normal 226 Hz tympanogram (peak-compensated static acoustic admittance between 0.3 and 2.5 mmhos and peak tympanometric pressure between −100 and +50 daPa).

Pure-tone air conduction thresholds were measured manually at octave and inter-octave frequencies between 0.25-8 kHz for each ear using a GSI Audiostar Pro audiometer (Grason-Stadler, Eden Prairie, MN) following the standard Hughson-Westlake procedure recommended by the American Speech-Language-Hearing Association (ASHA, 2005).

Pure-tone bone conduction thresholds were measured at octave frequencies from 0.5 to 4 kHz. Participants with a PTA greater than 55 dB were excluded to ensure stimuli presented at the fixed level of 65 dB SPL remained audible enough to avoid floor effects in performance. The ear with the better PTA (1, 2, and 4 kHz) was selected for testing. If the difference between ears was judged (by an audiologist) to be negligible, then the test ear was chosen randomly with a bias toward equalizing the number of right and left ears. Other definitions of PTA were explored but had minimal influence on a subset of results, so we had no compelling evidence to deviate from the definition that has often been used in previous studies in our lab. Participants were assigned to one of four hearing categories based on their audiometric PTA of the test ear for analysis. The categories were normal hearing (PTA-10 – 15 dB HL), slight hearing loss (16 – 25 dB HL), mild hearing loss (26 – 40 dB HL), and moderate hearing loss (41 – 55 dB HL). Table 1 shows the number of participants included in each study and reports hearing PTA category, sex, mean age, and age range.

**Table 1:** Participants included in each study, where n is the total number of participants. Pure-tone average (PTA 1, 2, 4 kHz) hearing category, sex (female (F) and male (M)), and mean age and age range (years) is reported. The hearing categories were normal hearing (NH; PTA-10 – 15 dB HL), slight hearing loss (16 – 25 dB HL), mild hearing loss (26 – 40 dB HL), and moderate hearing loss (41 – 55 dB HL).

| Participants | n | NH | Slight | Mild | Moderate | Sex | Age |
| --- | --- | --- | --- | --- | --- | --- | --- |
| Exp. 1 | 10 | 10 | 0 | 0 | 0 | 5 (F) 5 (M) | 29.6 (19-53) |
| Exp. 2 | 60 | 15 | 15 | 15 | 15 | 29 (F) 31 (M) | 55.5 (20-78) |
| Exp. 3 | 20 | 10 | 0 | 4 | 6 | 13 (F) 7 (M) | 46.1 (20-80) |
| Exp. 3 | 40 | 10 | 10 | 10 | 10 | 20 (F) 20 (M) | 54.5 (24-76) |

### Task Design

In Experiment 1, participants completed one practice block, consisting of one presentation each of 78 VCV tokens in quiet (3 vowels x 13 consonants x 2 recordings), and ten test blocks. Test blocks presented each of the 78 VCV tokens at three fixed SNRs: 0-, 6-, and 12-dB SNR. Each test block consisted of 234 randomized trials (3 vowels x 13 consonants × 2 recordings × 3 SNRs). Ten blocks were presented for 2340 total trials, which took an average of 95 minutes to complete, excluding breaks, across two lab visits.

There were two recordings of each VCV. Based on the similarity of the pairs of lines in Fig. 1, a judgment was made regarding the differences between the pairs of recordings as negligible. This assessment had little impact on the test procedures and could be revisited in a post hoc analysis of the test results. For vowel contexts, consonant recognition performance was most consistent across consonants for /ɑ/. The vowel contexts /i/ and /u/ showed greater variability and were largely collinear. The vowel context /u/ was removed from further consideration to mitigate collinearity and reduce variability.

**Figure 1.**
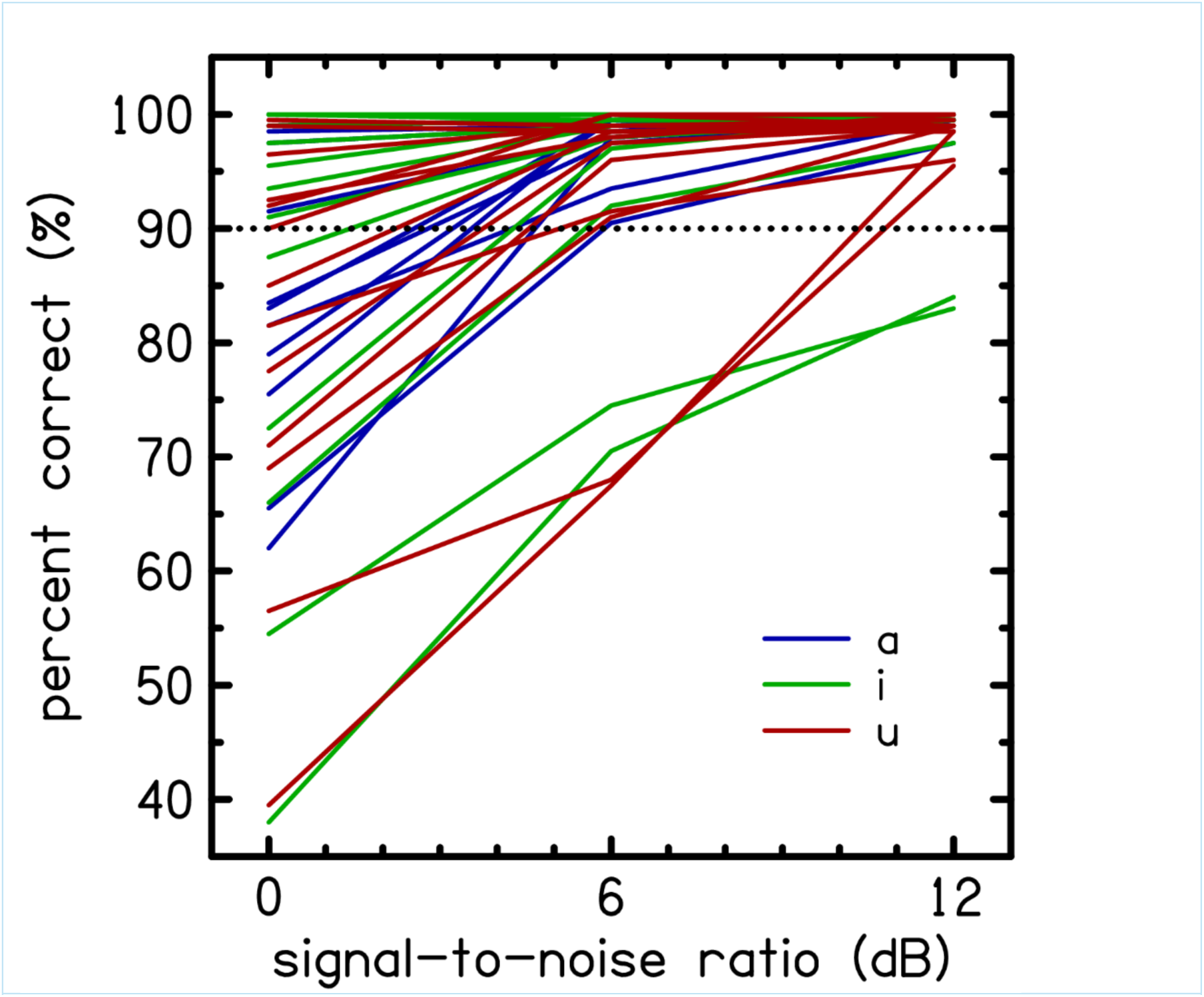
Consonant recognition performance in percent-correct for each consonant (unlabeled) and grouped by vowel context shown by color, /ɑ/ (blue), /i/ (green), and /u/ (red). For each fixed SNR (0, 6, 12 dB), VCV scores were averaged across participants in Experiment 1. The black dashed line indicates 90% performance.

For the remaining 26 VCVs, the curves in Fig. 1 were used to select a narrower range of SNRs that were likely to have a normal performance close to 90% (SNR90) for each VCV. Inspired by (Phatak et al., 2009), it was hypothesized that normalizing perceptual difficulty for the normal hearing group would cause those with hearing loss to fall on the steepest region of the performance intensity function. By normalizing in this manner, consonant confusions are more sensitive to hearing loss, making it possible to statistically differentiate between groups based on performance. Instead of measuring complete psychometric functions, which are costly and time-consuming, a narrow range of SNRs was extrapolated from a wider-spaced set of SNRs likely to encompass the SNR90. SNRs for each VCV were selected that were optimally sensitive to hearing loss by examining the AUC of each VCV. The ultimate justification for this selection was in the maintenance of sensitivity to hearing loss in the final set of stimulus tokens. Test performance variability was reduced further by removing the vowel context /u/ from the stimuli set, which dropped the total number of VCV tokens from 234 to 156 (2 vowels x 13 consonants x 2 recordings x 3 SNRs). Each VCV had three optimized SNRs (SNR90 +-3 dB). This was the stimuli set used in Experiment 2.

In Experiment 2,participants completed one practice block with 52 VCVs in quiet (2 vowels (/ɑ i/) x 13 consonants x 2 recordings), and ten test blocks. Each VCV token was remixed with three levels of speech-spectrum noise to achieve SNR90 (SNR90 ± 3 dB) specified by the values in Table 2, for a stimulus set that included 156 tokens (2 vowels x 13 consonants × 2 recordings × 3 SNRs). Tokens were randomized within each block for a total of 1560 trials, which took an average of 70 minutes to complete, excluding breaks, across two lab visits.

**Table 2.** Final ten consonants included in the qVCV test and their respective optimized signal-to-noise ratios (SNR) and area under the curve (AUC) values expressed as percent and rounded to the nearest integer.

| **Consonant** | **b** | **d** | **g** | **k** | **n** | **s** | **ʃ** | **t** | **v** | **z** |
| --- | --- | --- | --- | --- | --- | --- | --- | --- | --- | --- |
| SNR | 15 | 4 | 3 | 0 | 15 | 0 | 0 | 0 | 15 | 0 |
| AUC | 96 | 98 | 94 | 89 | 100 | 94 | 98 | 99 | 95 | 98 |

In Phase 2, the final qVCV test configuration was utilized. This set consisted of 10 specific consonants (/b d g p t k v z s ʃ m n/) presented in the /a/ vowel context. Unlike the Phase 1 Experiments, each consonant was presented at a specific, fixed SNR optimized to maximize sensitivity to hearing loss (see Table 2 for the specific SNR assigned to each consonant).

In Experiment 3, participants completed one practice block with 20 VCVs in quiet (1 vowel (/ɑ/) x 10 consonants x 2 recordings), and five test blocks. Tokens were presented at a single SNR (1 vowel x 10 consonants x 2 recordings x 1 SNR) and repeated five times each in a test block, totaling 100 trials per block. Tokens were randomized within each block for a total of 500 trials, which took an average of 20 minutes to complete, excluding breaks, in a single lab visit. All participants completed the qVCV test in the unaided condition and the participants who had hearing loss also repeated the protocol in an aided condition, where they wore their personal hearing aid in its default setting underneath the circumaural headphone (Sennheiser HD 280 Pro). The audiologist confirmed that the hearing aid was powered on and functioning via a listening check, but no changes to the settings or prescriptions were made. Aided testing took an additional 20 minutes and was completed during the same lab visit.

In Experiment 4, the same set of 20 VCV tokens (10 VCVs x 2 recordings x 1 SNR) was used. All participants completed one practice block in quiet, and then two test blocks consisting of five repetitions of each of the 20 VCVs (100 trials per block) for a total of 200 trials, which took an average of 10 minutes to complete in a single lab visit. All participants completed the testing in the unaided condition (n=40), and participants in the mild and moderate hearing loss categories (n=20) repeated the tests with a hearing aid fit by an audiologist in the laboratory and with a personal hearing aid if they had one (n=9). The lab hearing aid was a commercial device fitted by an Audiologist to NAL-NL2 prescriptive targets for each participant using the Verifit 2 system (Audioscan, Ontario). Real-ear measures were obtained with personal hearing aids, but no changes were made to device programming or settings. The hearing aid was worn on the ear underneath the circumaural headphone, and the participant completed one practice block in quiet followed by one test block of 100 trials (five repetitions each of the 20 VCVs) in the same lab visit.

The QuickSIN test was also administered in Experiment 4. Two list pairs (four lists in total; Lists 1, 8 and 6, 15) were selected from the QuickSIN, which were shown to have equivalent reliability (*QuickSIN Speech-in-Noise Test User Manual*, 2001). The lists were presented monoaurally at 70 dB sound pressure level (SPL). The SNR was 25 dB for the first sentence and decreased in 5 dB steps until it reached 0 dB for the last sentence. Participants were given instructions based on the QuickSIN user manual. A practice list was first administered (Practice List A/21) and the participant was allowed to ask questions if any further instructions were needed. Then, four lists were administered (in randomized order) and the tester live-scored key words as correct or incorrect.

All participants completed the QuickSIN protocol in the unaided condition, and participants in the mild and moderate HL categories also completed testing in aided conditions following the procedures described above. Different lists were used and only one list-pair (two lists) was administered in the lab-fit HA condition (Practice List B, lists 10 and 11) and personal HA condition (Practice List C, lists 2 and 12). The presentation order of qVCV and QuickSIN in Experiment 4 was counterbalanced across participants to control for test order effects. The sequence order of the aided conditions, lab-fit HA and personal HA, was also counterbalanced across participants.

## Results

### Phase 1

Experiment 1 data were used to improve the VCV test by reducing variability in the vowel context and to determine optimal SNRs for VCVs that more precisely estimate a SNR range likely to normalize recognition performance close to 90% for normal hearing listeners. Figure 1 shows the consonant recognition performance, averaged across participants, as a percentage correct for each of the 39 VCVs as a function of SNR. Tokens were grouped by vowel context: /ɑ/ in blue, /i/ in green, and /u/ in red. Of the 78 VCV tokens, 63 intercepted the 90% target between 0 and 6 dB SNR. However, Fig. 1 illustrates that the 0-12 dB SNR range was insufficient for some VCVs. For example, the lowest green lines correspond to /*ibi*/ and /*imi*/ and for the highest SNR of 12 dB, the average performances were 84% and 83%, respectively. Performance saturated with nearly 100% recognition rates for the consonants /*ʃ t z*/ for all vowel contexts and /*g s*/ for /*ɑ i*/ and /*ɑkɑ*/.

Logistic regressions were used on the Experiment 2 data to determine which VCVs performed best at separating ears with normal hearing from those with hearing loss, defined as PTA greater than 15 dB HL. The primary objective of this experiment was to separate these two categories. Binomial logistic regression on the principal components of the VCV data was used to separate the ears without hearing loss from those with hearing loss. Following cross-validation, the model made a binary prediction of hearing loss. The area under the curve (AUC) was used to evaluate the discrimination power of each VCV or a subset of VCVs to predict hearing loss. AUC values between ≥0.7 are commonly used to denote clinical significance. The AUC of each vowel /ɑ/ or /i/ using all 13 consonants was used to evaluate which of the two vowels had better predictive performance.

The predictive likelihood of each VCV token to separate participants with normal hearing from those with hearing loss is expressed as AUC in Figure 2, which shows the AUC distribution for each consonant. /f/ and /m/ had a relatively large distribution of AUC values across participants, and the consonants /f m p/ had the lowest AUCs (below 0.9). Therefore, /f m p/ were removed from subsequent analyses and experiments.

**Figure 2.**
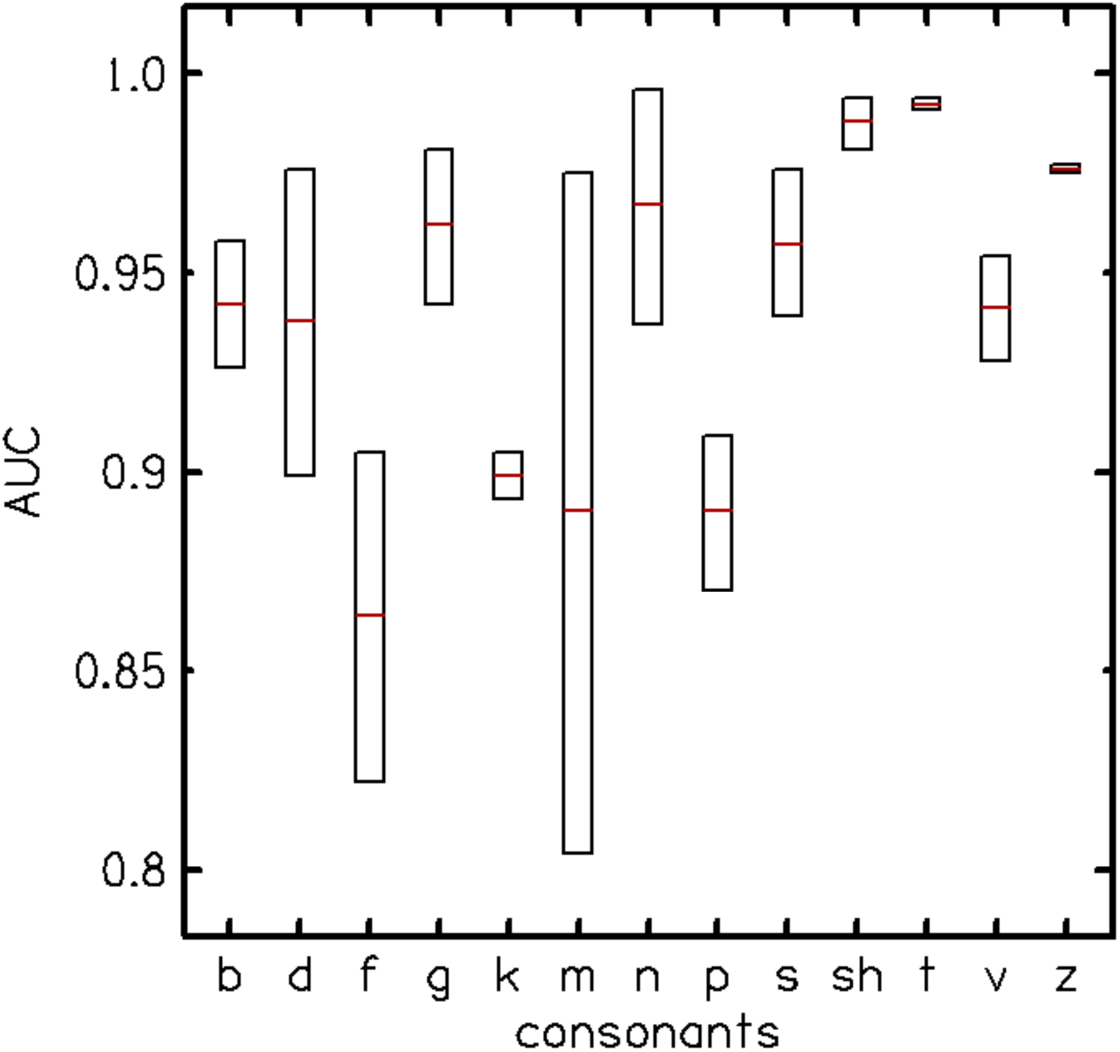
Area under the curve (AUC) distribution across vowels for each consonant shows the predictive likelihood of each consonant for separating participants with normal hearing from those with hearing loss. The consonants /f m p/ had the lowest test performance.

After removing the three poorest-performing consonants, the AUCs for each vowel context were reanalyzed, as shown in Figure 3. Fig. 3 suggests that the vowel context /ɑ/ is optimal because it has less variance and a higher median AUC (∼0.97) than /i/, which has a median AUC of ∼0.95. The AUC values also show that only one vowel context is required to separate normal hearing from hearing loss ears; hence, the /i/-vowel context was removed from the stimulus set. As detailed in the Methods, the optimization process resulted in a final set of 10 VCVs in the vowel context /a/, each assigned a single SNR (Table 2). This selection achieved the highest AUC for separating normal hearing from hearing-impaired ears. Figure 4 shows the performance of the 60 participants in the Experiment 2 as an average score of the qVCV stimuli trials in percentage correct as a function of their measured audiometric PTA. The percent correct score was computed as the sum of the diagonal elements of the consonant confusion matrix (CCM).

**Figure 3.**
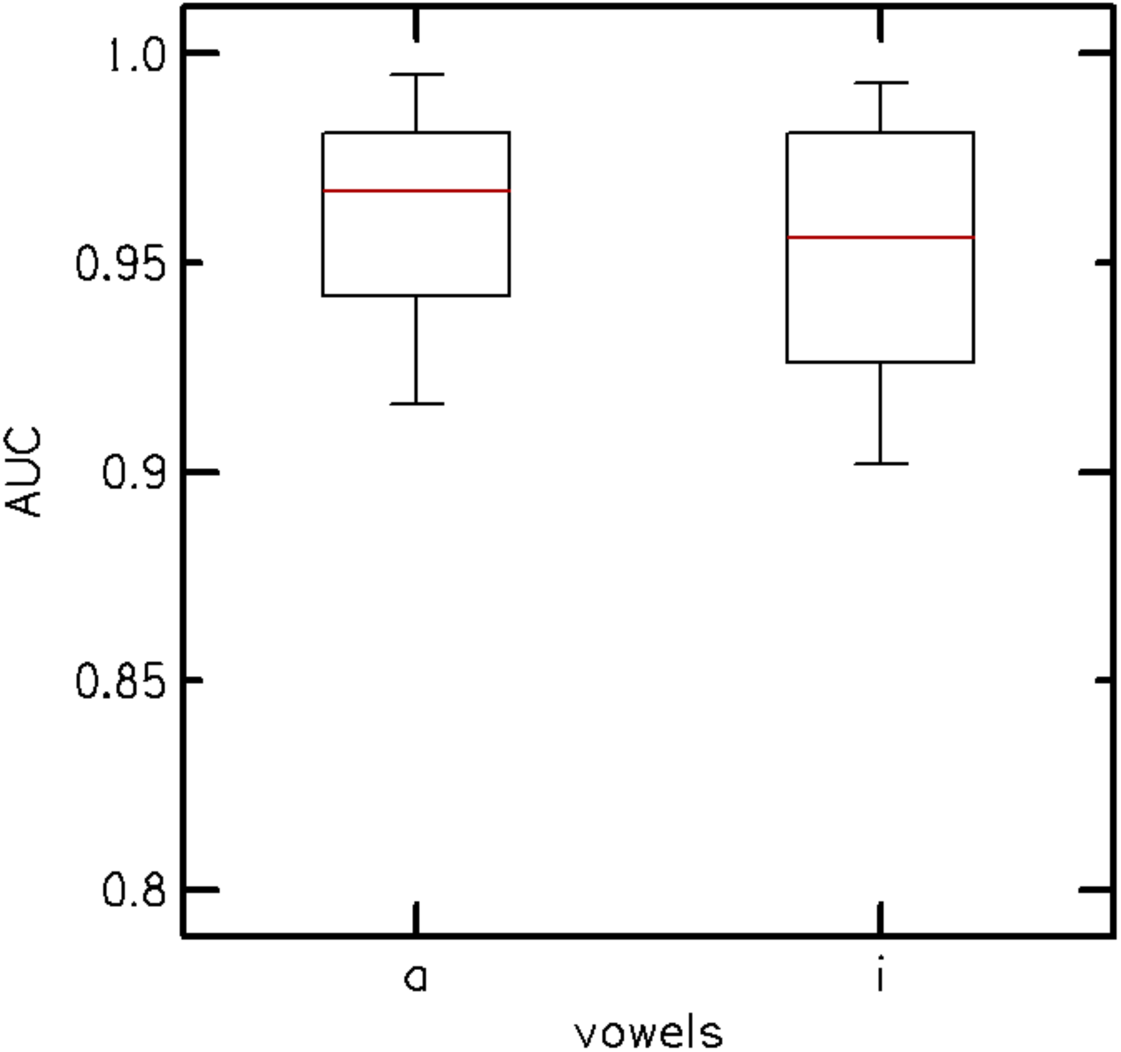
The area under the curve (AUC) distribution for vowel contexts /ɑ/ and /i/ after removing /f m p/ from the analysis. The vowel context /ɑ/ has less variance and a higher AUC (0.97) relative to /i/ (0.95).

**Figure 4.**
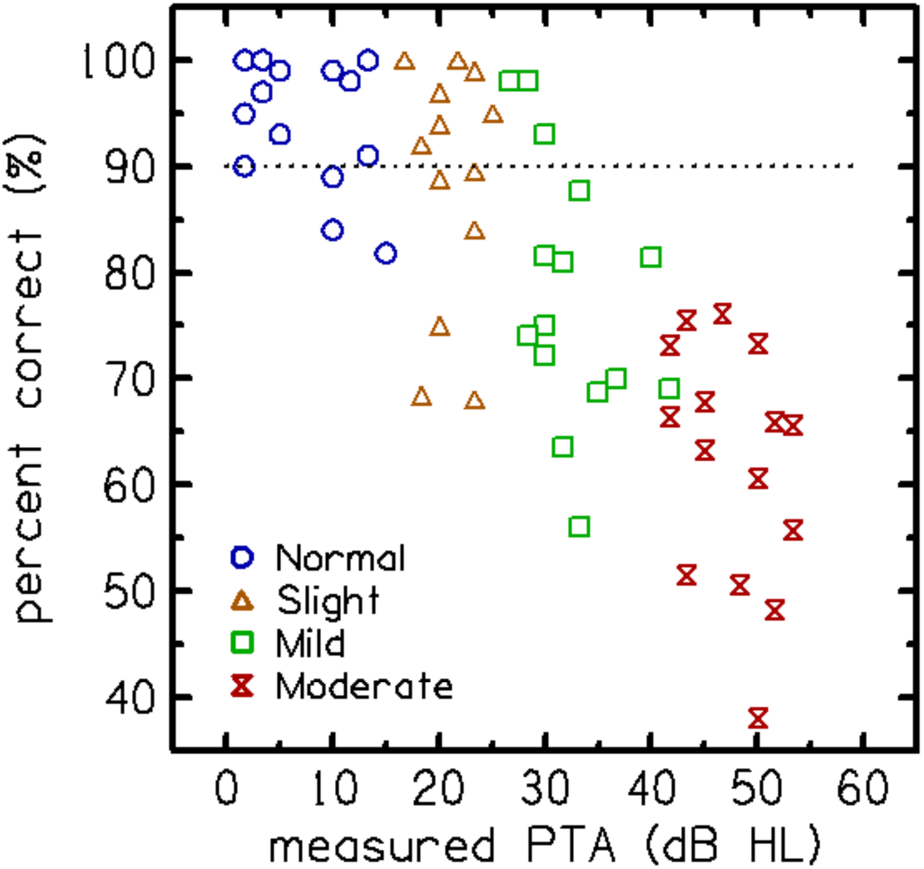
Individual consonant recognition performance (in percent-correct) on the qVCV test as a function of measured audiometric pure-tone average (PTA). Colored symbols denote the HL category to which participants from Experiment 2 were assigned, as defined in Table 1.

To predict PTA from the qVCV performance, we utilized a logistic regression model. First, the Consonant Confusion Matrices (CCMs) were vectorized. Principal Component Analysis (PCA) was then performed on these vectors to reduce dimensionality and minimize multicollinearity among the confusion patterns. The resulting principal components were used as the predictor variables in the regularized logistic regression to compute the ‘predicted PTA’ estimates. These regressions implemented regularization and cross-validation (utilizing 70% for training and 30% for validation) to reduce overfitting and to generalize the prediction performance. A regularization factor, λ = 3, was used, which minimized prediction error in the validation set. Figure 5 shows the predicted PTA on the y-axis and the actual, measured audiometric PTA on the x-axis. The output of the logistic regression model showed that PTA predictions for the hearing loss groups fell along the diagonal. The “audibility function” (dashed line in panel b), fitted to the predicted PTA, has a unity slope for ears with measured PTA above 15 dB HL and is constant (15 dB HL) for ears with lower measured PTA. The cross-validated prediction of the individual PTA had a mean absolute error of 5.7 dB.

**Figure 5.**
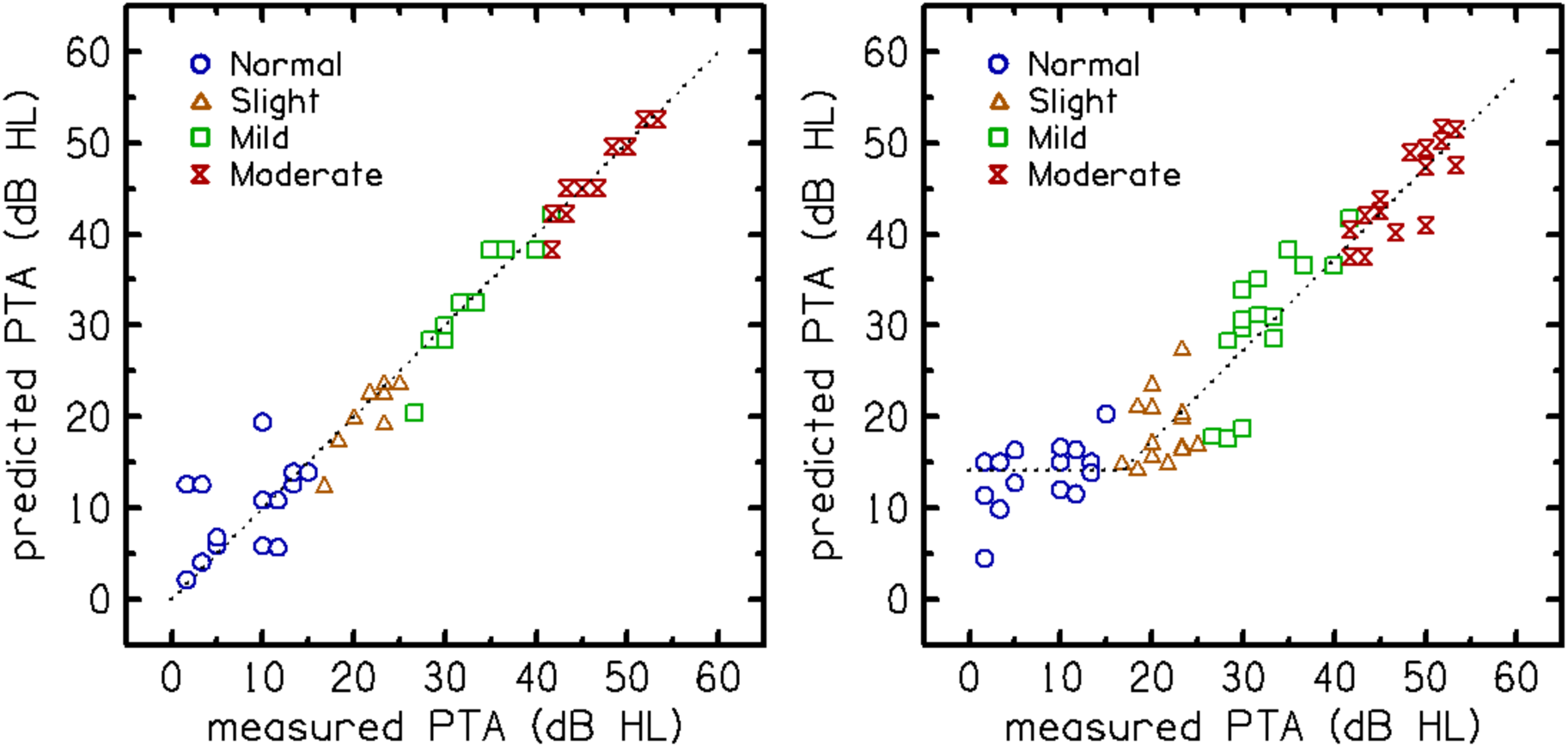
Predictions of pure-tone average (PTA) from qVCV scores and a logistic regression model. Each point represents an individual participant’s (Experiment 2) predicted PTA (y-axis) plotted against their measured audiometric PTA (x-axis). Participants are grouped (colored symbol) according to the degree of hearing loss. Panel a (left) shows PTA prediction without cross-validation (training performance), illustrating potential overfitting. Panel b (right) shows PTA predictions after cross-validation (validation performance), representing the model’s generalization to new data. The audibility function (dashed line in panel b) is a piecewise baseline fitted to the cross-validated predictions to account for the floor effect in normal hearing. It has a unity slope for ears with measured PTA above 15 dB HL and is constant (15 dB HL) for ears with lower measured PTA. This function serves as the baseline for the residual loss calculation in Figure 6.

The “audibility function” fitted to the predicted PTA, shown as a dashed line in Fig. 5b, was subtracted from the predicted PTA to obtain the “residual loss”, as illustrated in Figure 6. Seven participants were identified as likely to have “excess loss,” which was defined as having residual hearing loss greater than 5 dB HL. This threshold was selected because it exceeds the test-retest repeatability limit of the qVCV (approximately 5 dB) and is nearly double the mean absolute error of the residual loss estimate (2.95 dB), ensuring that identified deviations are unlikely to result from measurement error alone. Five of these seven participants were over 60 years old, five were male, four had a history of noise exposure, and six self-reported bilateral tinnitus. The mean absolute error for the estimated residual hearing losses was calculated to be 2.95 dB, with an AUC of 0.76, which is clinically acceptable.

**Figure 6:**
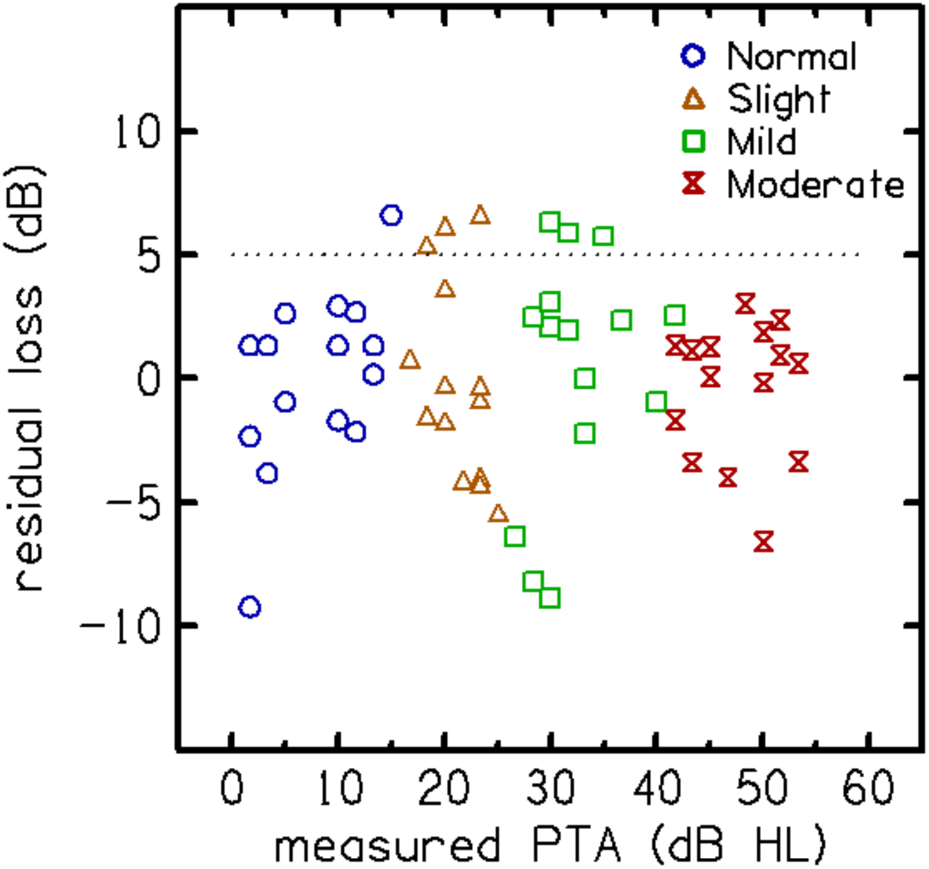
Residual hearing loss (RHL) was computed by subtracting audibility (dashed line in Fig. 5b) from the measured audiometric pure-tone average (PTA). Individual estimates of RHL are shown as a function of measured PTA. Seven participants in Experiment 2 had RHL > 5 dB (dashed line).

### Phase 2

Experiment 3: The expected performance of the qVCV test in quantifying HA benefit was verified in a new set of 20 participants (10 NH and 10 HL). Participants with NH scored better than expected on the test (not shown), near ceiling. In participants with hearing loss, performance improved with a hearing aid compared to without a hearing aid, but performance was not restored to that of peers with normal hearing. The HA benefit was lower for participants with lower PTAs.

Experiment 4: With evidence that the qVCV test showed potential for quantifying HA benefit in the laboratory, it was important to measure its test repeatability and eeiciency compared to a SIN test that has been adopted into common clinical practice to assess the clinical viability of the qVCV test. Figure 7 (left panel) shows the relationship between the percentage correct scores on qVCV and audiometric PTA. Participants with normal hearing scored above 90% correct on qVCV, and the scores systematically declined as the degree of hearing loss increased. The relationship between actual and predicted PTA is shown in the right panel. The audibility function fitted to the PPTA, shown as a dashed line, was constant (at 15 dB HL) for ears with an actual PTA below 20 dB HL (i.e., normal hearing) and equal to the PTA for ears with hearing loss (actual PTA above 20 dB HL). The predicted PTA systematically increased as audiometric PTA increased in participants with hearing loss.

**Figure 7.**
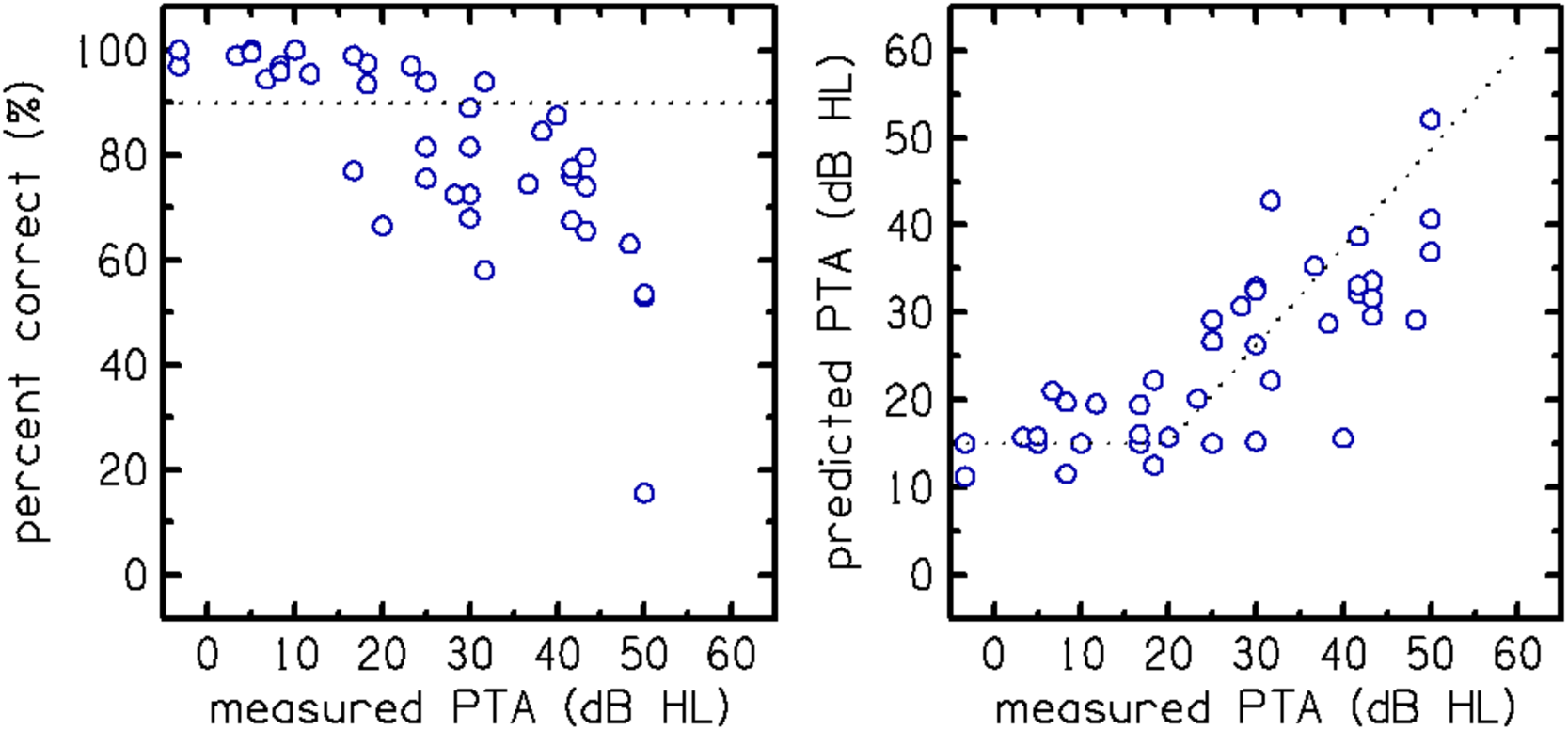
The relationships between qVCV scores and audiometric pure-tone average (PTA) are shown in percent-correct (left panel) and predicted PTA (right panel) for Experiment 4. Each data point represents an individual participant’s consonant recognition performance as a function of their measured PTA. The audibility function (right panel; dashed line) fitted to the predicted PTA is constant (15 dB HL) for normal hearing ears (PTA < 20 dB HL) and equal to the PTA for hearing loss ears (PTA > 20 dB HL).

Test-retest repeatability was evaluated for both tests by comparing scores between repeated tests. Figure 8 shows the QuickSIN scores (top row) in percent-correct scores (left panel) and SNR loss scores (right panel) for test 1 vs. test 2. The correlation coefficients between the two scores were both R=0.945, indicating good repeatability between the first and second tests. The qVCV scores (bottom row) for percent-correct scores (left panel) and PPTA scores (right panel), also showed good repeatability with correlation coefficients between the two tests of R=0.974 and R=0.907, respectively. This demonstrates that QuickSIN and qVCV had similar repeatability.

**Figure 8.**
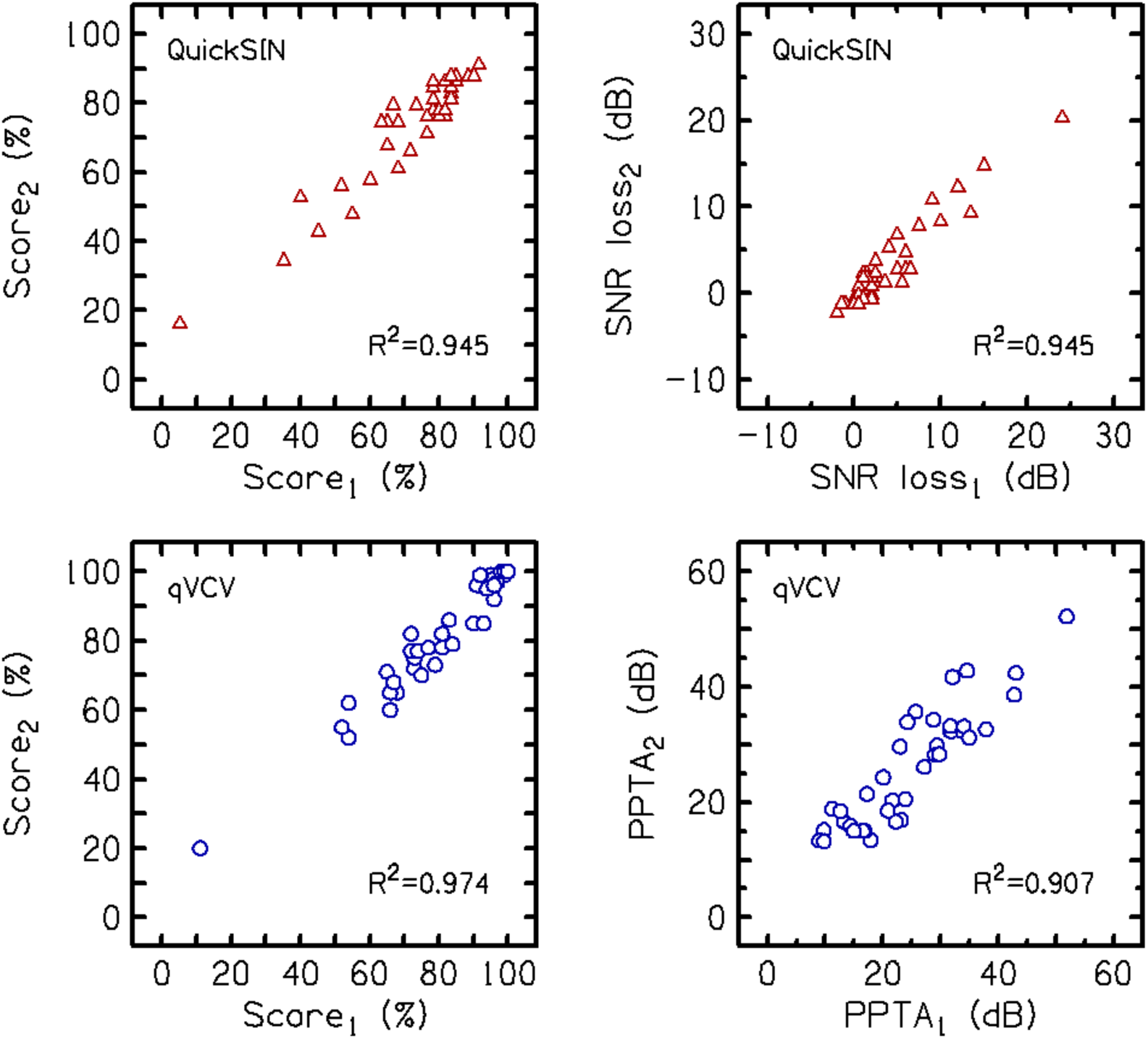
Comparison of scores between repeated trials on the QuickSIN (top row) and the qVCV (bottom row) in Experiment 4. Scores are shown as percent-correct (left column) and hearing loss measures (right column).

To assess clinical efficiency, we examined how test repeatability (defined as the Mean Absolute Difference between sessions) improved as the test duration increased (Figure 9). The analysis revealed that the qVCV achieves stable repeatability, comparable to the average of two QuickSIN lists, after approximately 50 trials. Given a presentation rate of 25 trials per minute, this indicates that a reliable score can be obtained in just two minutes.

**Figure 9.**
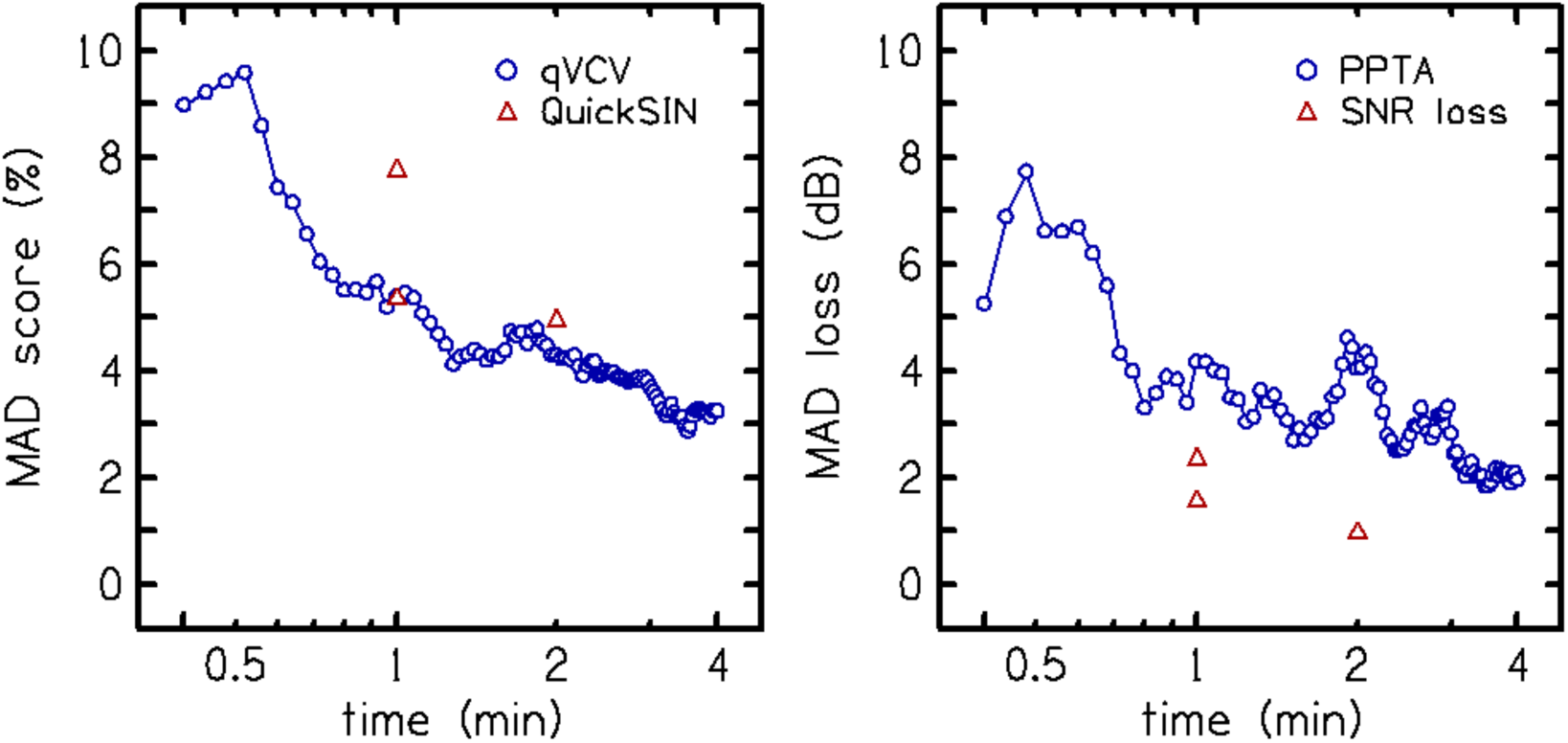
Mean-absolute differences (MAD) between test sessions are shown as a function of test time (minutes) for percent-correct (left panel) and hearing loss measures (PPTA for qVCV and SNR loss for QuickSIN; right panel). qVCV test time assumes 25 trials per minute and one QuickSIN list assumes one minute. QuickSIN test times for one and two lists are placed at one and two minutes, respectively.

Although the qVCV and QuickSIN tests are not identical measures of speech-in-noise performance, given the similarity between the test reliability and outcome measures, it is possible to compare the relationship of scores between the tests. Figure 10 shows the correlations between the test performance in percentage correct (left panel) and dB hearing loss measures (PPTA for qVCV and SNR loss for QuickSIN; right panel). The correlations were R^2^=0.732 for percent-correct scores and R^2^=0.617 for estimated hearing loss measures (in dB).

**Figure 10.**
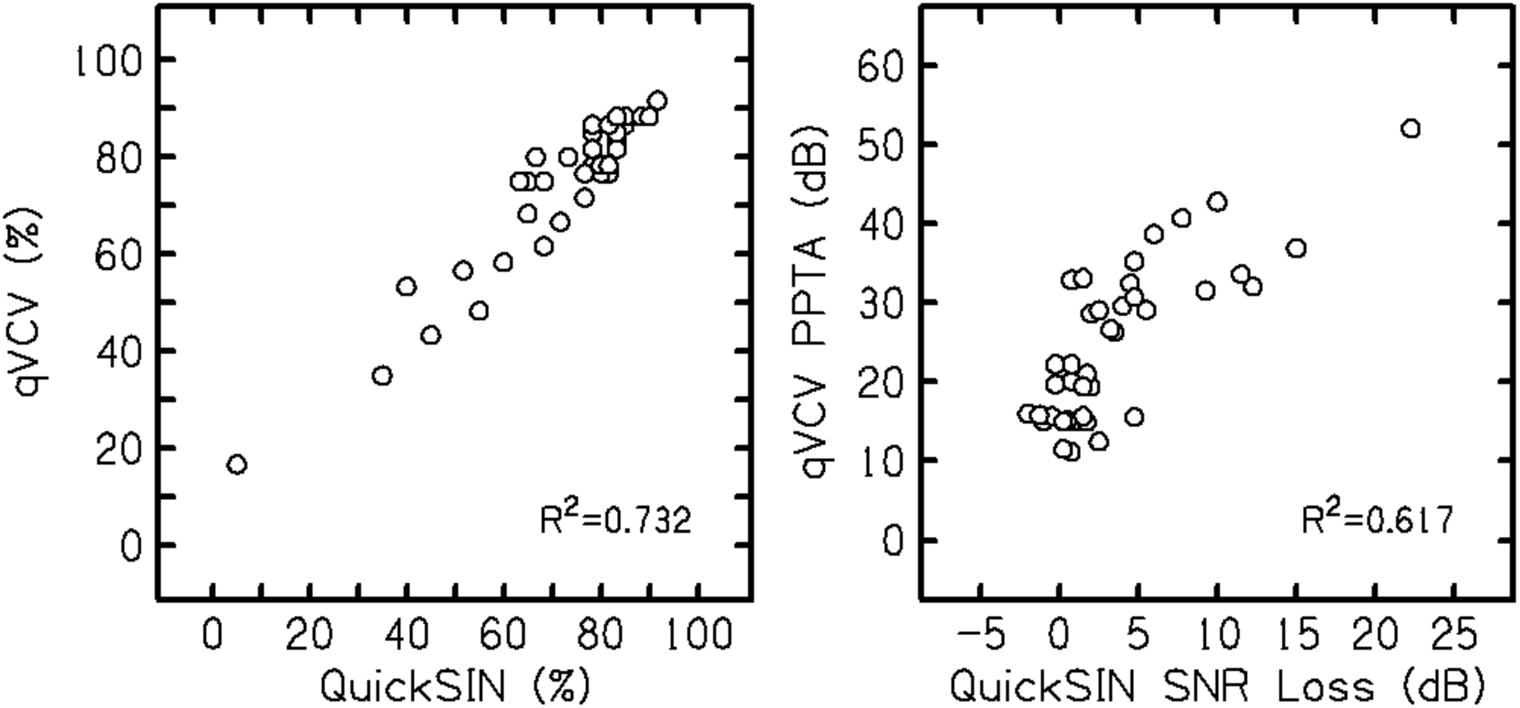
QuickSIN test results (x-axis) compared to qVCV test results (y-axis) for percent-correct scores (left panel) and hearing loss measures (PPTA for qVCV and SNR loss for QuickSIN; right panel) from Experiment 4.

Figure 11 shows hearing aid benefit (HAB; difference between aided and unaided scores) as a function of the measured audiometric PTA. HAB significance is defined as dB improvement above the repeatability limits for each test (2.5 and 5 dB for QuickSIN and qVCV), indicated by the dashed lines in the bottom panels. HAB was observed in both tests; however, QuickSIN did not indicate HAB for ears with PTA <40 dB and qVCV reflected benefit across a wider range of PTA. Figure 11 illustrates the magnitude of the hearing aid benefit (difference scores); however, analysis of the absolute scores revealed that neither lab-fit nor personal hearing aids restored performance to the ceiling levels (typically >90%) observed in NH participants. This suggests that these hearing aids did not typically restore normal consonant recognition.

**Figure 11.**
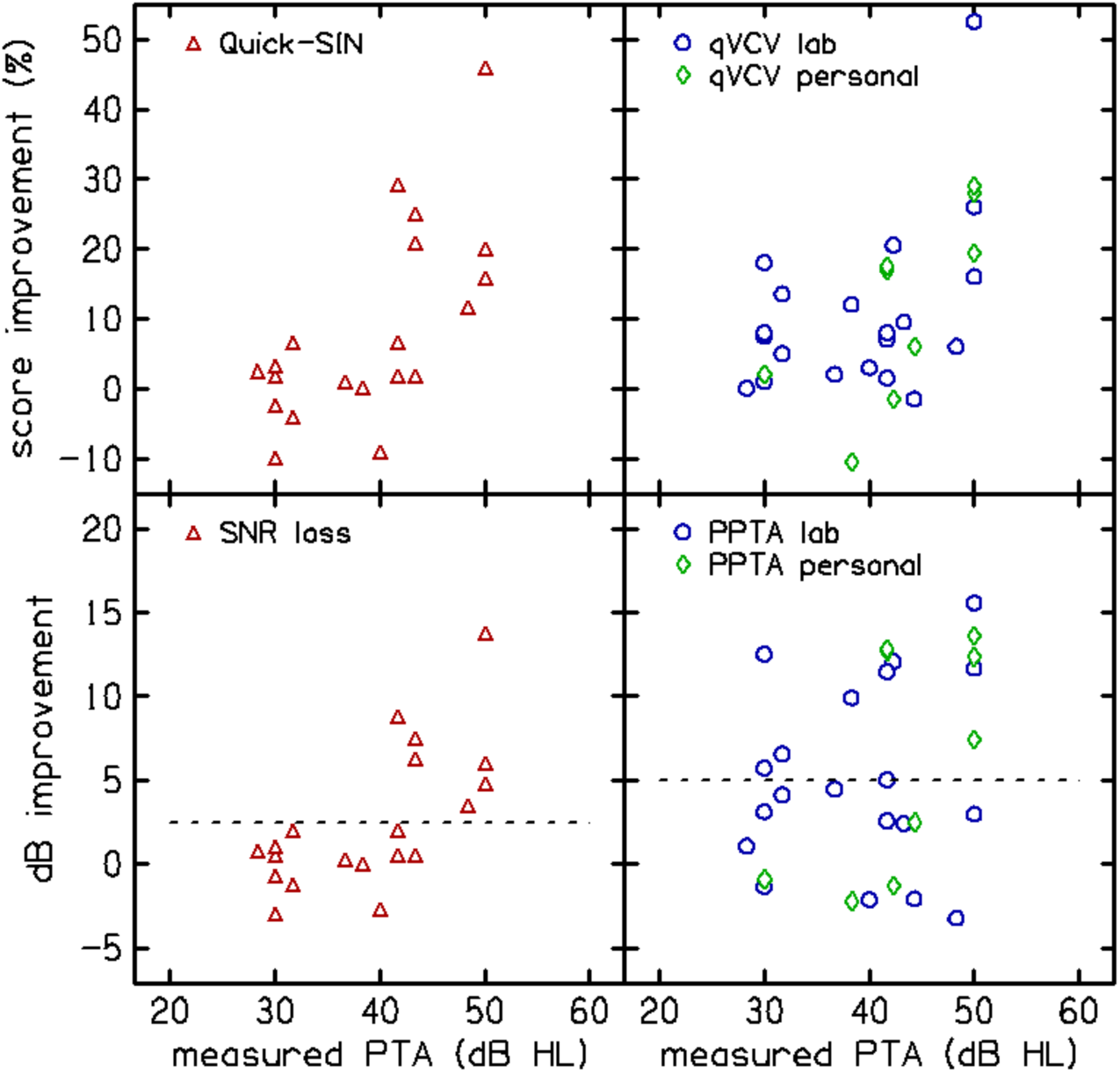
Hearing aid benefit (HAB; difference between aided and unaided scores) shown as a function of measured audiometric pure-tone average (PTA). QuickSIN scores are shown in the left column; qVCV scores are in the right column. Percent-correct improvement is shown in the top row; dB improvement is shown in the bottom row.

The Speech Intelligibility Index (SII) was used as a measure of device fit and was compared to HAB (qVCV PPTA dB improvement between the unaided and aided conditions). The Verifit 2 was used to estimate unaided SIIs for each participant based on the audiogram (250 – 8 kHz) and real-ear measurements were recorded for lab and personal hearing aids at 65 dB SPL to estimate aided SII. he aided SII was significantly higher than the unaided SII for all participants. However, despite the strong dependence of the qVCV on audibility, linear regression analyses revealed no significant relationship between the Hearing Aid Benefit (HAB) measured by qVCV and the SII (analyzed as unaided SII, aided SII, or the difference between them). This suggests that while the hearing aids restored spectral audibility (as measured by SII), this improvement did not linearly translate into improved consonant resolution for all listeners, likely due to suprathreshold deficits not captured by the SII. The lab-aided SII was higher than the personal-aided SII for 7/8 participants. However, there was no significant relationship between the device and HAB.

## Discussion

Audiologists require efficient tools that provide useful information about the patient to make clinical decisions. The audiogram alone does not always explain a patient’s self-perceived hearing difficulties or the benefit they may or may not receive from a hearing aid device. Many speech-in-noise tests exist that assess an individual’s performance of speech perception in background noise (e.g., DIN, HINT, QuickSIN, WIN, among others), but each has its own limitations, including concerns regarding test time, individual variability, cognitive demands, and scoring and interpretation of results.

Most speech recognition errors manifest as consonant confusions, which occurs when a listener perceives a spoken consonant as a different consonant (Miller & Nicely, 1955; Phatak et al., 2009; Phatak & Allen, 2007; Wang & Bilger, 1973). Consonant confusion analysis takes advantage of the ability to examine audibility within frequency bands to predict hearing status and listening difficulty in ways that are not possible with other types of speech stimuli. Masking consonants with noise and measuring confusion matrices results in a wide range of variability across consonants, listeners, and hearing configurations (e.g., (Dubno et al., 1982; Gordon-Salant, 1985; Miller & Nicely, 1955; Wang & Bilger, 1973). Phatak et al., (2009) used noise to mask vowels, consonants, and consonant-vowels with different signal-to-noise ratios (SNR). Their results showed that averaging across vowel contexts increases variability. One strategy to minimize variability would be to test consonants in a single-vowel context. Furthermore, presenting a single stimulus per trial, rather than multiple words or sentences, and using a forced-choice task from a closed set may reduce the influence on performance of working memory and cognitive load (Toscano & Allen, 2014).

Our goal was to develop a quick, multifaceted speech-in-noise task that is not only sensitive to hearing loss but also has predictive capabilities for assessing hearing aid benefit. The development of the qVCV test progressed across two phases of experimental testing. Although many details layer the development of the qVCV test, the ultimate justification for this process lies in the result, which is a low-cognitive-load speech-in-noise test with demonstrated efficiency, repeatability, and sensitivity to hearing loss. We acknowledge that a full confusion matrix is diagnostically rich. However, the specific aim of the qVCV test is clinical efficiency and the derivation of a metric (Predicted PTA) rather than a phoneme-level diagnostic profile. By selecting only the most sensitive tokens, we reduce test time to approximately 5 minutes, making it viable for routine clinical use. This trade-off was intentional to prioritize sensitivity to overall hearing status over specific phonemic diagnostics.

It is important to note that because the stimuli are presented at a fixed level of 65 dB SPL, the raw percent-correct score is heavily influenced by hearing loss. This is reflected in the strong correlation between qVCV scores and audiometric PTA. However, the qVCV approach is distinctly different from other speech tests that hold the performance target, typically threshold, constant while varying the stimulus level to determine the influence of hearing loss. The qVCV test, in its current form, is optimized for mild-to-moderate loss profiles, where it is more likely to capture perceived hearing deficits that are greater than expected based on the audiogram alone and where recommendation for hearing aids isn’t always straightforward.

An examination of residual loss, defined as the difference between the actual and predicted PTA, identified seven listeners in Experiment 2 as likely to have greater-than-expected loss (Fig. 6). A regression analysis suggested that the presence of excessive loss, defined in this study as having residual loss greater than 5 dB, could be predicted, even without prior knowledge of the actual PTA, with AUC=0.76, which is clinically acceptable (see Fig. 5). Figure 6 shows that some listeners had negative residual losses, indicating performance that was better than predicted. Most of these values fell within the ±5 dB test-retest variability limit established for the qVCV, suggesting they are primarily attributable to random measurement error rather than a systematic physiological mechanism. Fitzgerald et al. (2023) observed a similar distribution of ‘better-than-predicted’ scores using the QuickSIN.

Table 3 summarizes the demographics of listeners with normal hearing (NH), hearing loss (HL), and those predicted to have excessive loss (XL). Hearing status was classified dichotically here by audiometric PTA ≤ 20 dB HL (NH) or PTA > 20 dB HL (HL), rather than into four categories, to reflect a screening paradigm that catches mild hearing losses. The excess loss (XL) group had RHL ≥ 5 dB. The table also includes the prevalence of self-reported hyperacusis, noise exposure, and bilateral or unilateral tinnitus, which were similar across participants in both experiments, and modified-SSQ score for Experiment 4. A potential translational benefit of predicting PTA using a clinically feasible consonant confusion test is its potential to quantify unexplained, or greater-than-expected hearing difficulties. However, the participants in the XL group in Experiment 4 did not all have elevated SSQ scores as expected. The scores ranged from 1.83-9.67 with a mean of 5.87, which was not different from the HL group. One participant in the XL category did not report any tinnitus, hyperacusis, or noise exposure and did not report any self-perceived difficulties in the SSQ. Further analysis of individuals who self-report hearing difficulties in their daily lives and who are identified with excess loss in qVCV is needed to explore the presence of underlying factors.

**Table 3.**
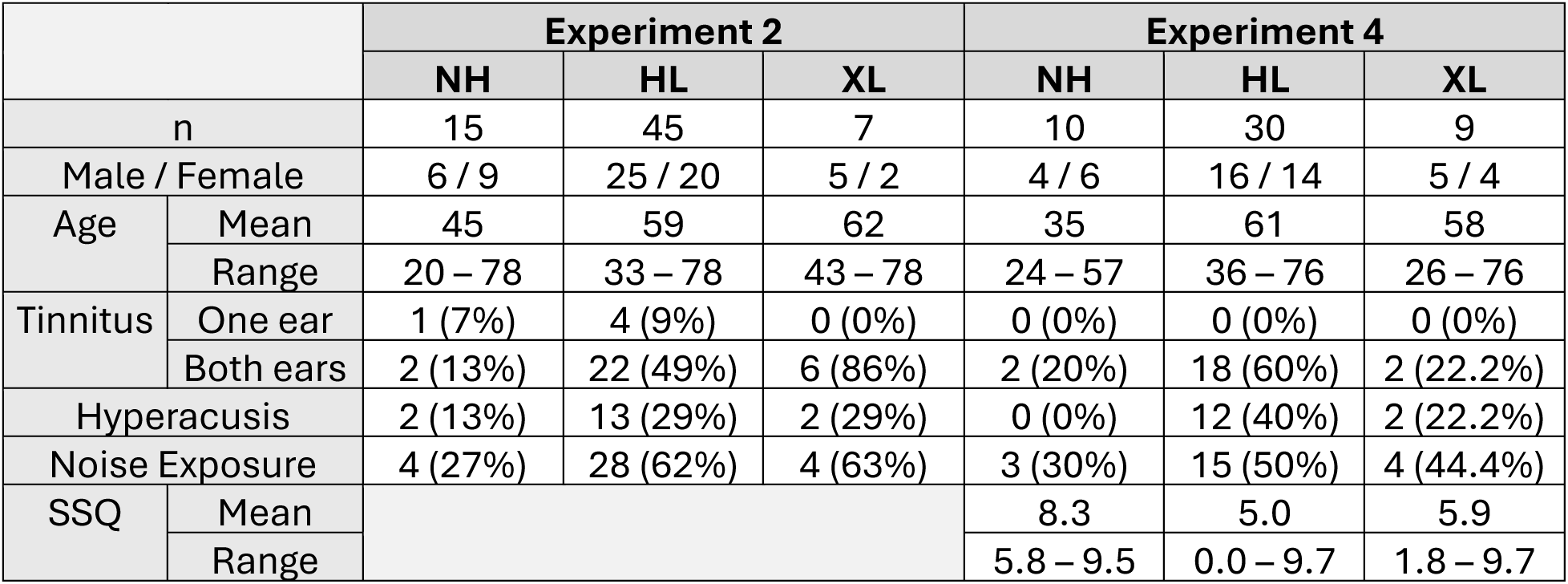
Summary of participant demographics including self-reported tinnitus, hyperacusis, and noise exposure for Experiments 2 and 4, and modified-SSQ score for Experiment 4. Participants are grouped by audiometric PTA ≤ 20 dB HL (normal hearing; NH), PTA > 20 dB HL (hearing loss; HL), and RHL > 5 dB (excess loss; XL). The participants in the XL group were also included in the NH and HL groups. The values in parentheses indicate within-group percentages. The sex and age distributions were similar between Experiments, as was the prevalence of tinnitus, hyperacusis, and history of noise exposure, which was higher in the HL groups than in the NH groups but lower in the Experiment 4 XL group than in the Experiment 2 XL group.

It is important to know how the qVCV test qualities compare to the properties of other currently employed clinical speech-in-noise tests before recommending its use. The qVCV test is as repeatable and efficient as the QuickSIN and can be administered in approximately 5 minutes. In addition to assessing the quality of the tests themselves, we can compare the relationship between test scores from subjects who completed both tests because the test outcomes (percent-correct and estimated hearing loss scores) are similar measures. Participants with more hearing loss showed greater loss on both qVCV and QuickSIN.

In Experiment 4, several participants reported that QuickSIN caused them to fatigue faster and stressed more because they worked hard to concentrate on the words being presented. One participant stated feeling that their memory was being tested and not their hearing; consistent with reports of feeling overwhelmed or discouraged over the test and that listening effort for different tasks is likely multidimensional (Alhanbali et al., 2019; Larsby et al., 2008; Pichora-Fuller et al., 2016). Qualitatively, when participants were asked about their experience during the qVCV test, the majority stated that the overall task was easy to comprehend and they felt they had more control over the speed at which the material was being presented. Unlike some other commonly employed speech-in-noise tests, one benefit of the qVCV test and our testing approach by design is to reduce reliance on working memory and cognitive load. Based on participant feedback, the qVCV test may be better accepted than the QuickSIN by patients in clinical settings.

Considering these advantages, another valuable application of qVCV may be the evaluation of hearing aid benefit. The clinical need for a quick, low-demand, test of hearing aid benefit was the driving force for the development of the qVCV test. Because the predicted PTA obtained in an unaided condition can indicate hearing loss, it may also support a recommendation for amplification. The qVCV test can be administered in aided conditions where aided benefit can be quantified and aided benefit between multiple devices, prescriptions, signal processing strategies, and settings can be compared which may lead to improved clinical outcomes and satisfaction.

Consonant confusion patterns have been linked to specific hearing loss configurations (Dubno et al., 1982; Phatak et al., 2009). A possible limitation of our analysis is that we observed several audiogram subtypes within our hearing loss groups, which may not have been adequately represented using the three-frequency PTA, which was the average of the pure tone thresholds at 1, 2, and 4 kHz. An alternative regression model could be developed using the same methodology presented here but with a dieerent definition of PTA or by replacing the predicted and measured PTA with the audibility index or speech intelligibility index (*Methods for the Calculation of the Speech Intelligibility Index*, 1997). Further exploration of consonant confusion matrices generated from the qVCV test in future studies may reveal other relationships across various hearing loss configurations.

A primary limitation of the hearing aid benefit assessment in this study was the presentation of stimuli via circumaural headphones placed over the hearing aids, rather than free-field presentation. While this method allows for ear-specific testing and eliminates room acoustic variables, it effectively alters the frequency response of the hearing aid and carries a risk of feedback. Feedback was monitored via listening checks, and participants used their personal devices in ‘default’ settings, which may include feedback suppression artifacts when coupled with headphones. While our results showed measurable benefit, clinical validation of the qVCV should ideally be conducted in a sound field environment.

We emphasize that the methods we applied to make the qVCV test sensitive to hearing loss can be adapted to create stimulus sets or tests responsive to effects other than hearing loss. The general procedure would be to 1) decide what variable (condition or effect) you want the test to be sensitive to, 2) place the variable of interest on the x-axis of your psychometric function, and 3) adjust your stimulus so that your control group achieves approximately 90% performance. The qVCV test software, including the stimuli and MATLAB scripts for analyzing the acquired consonant confusions with our logistic regression models, as well as a User Manual, is available in a GitHub repository (*qVCV Test*, BTNRH [Software]. GitHub).

## Conclusions

The set of ten consonants in a single vowel context selected for the qVCV test has been shown to be sensitive to hearing loss in a clinically feasible amount of time. To our knowledge, the ability of qVCV to predict pure-tone average (PTA) thresholds within 5.6 dB is unprecedented. Compared to QuickSIN, qVCV has several advantages, including lower cognitive demand, fewer learning effects, and automated scoring. qVCV may be used as an objective clinical tool to identify individuals with greater-than-expected hearing difficulties based on their audiogram alone, which is consistent with hypotheses regarding cochlear neural degeneration or cognitive deficits, though further physiological verification is required. Predicted PTA may also provide clinical feedback to patients regarding the benefit they receive from their hearing aid.

## Data Availability

All data produced in the present study are available upon reasonable request to the authors

## Acknowledgments

This research was supported by NIH grants R01-DC0008318 and P20-GM109023. We thank Aryn Kamerer and Laurencia Santillan for their contributions to the data collection and preliminary reports. Portions of this paper were presented as posters at the 2018, 2022, and 2024 American Auditory Society Annual Scientific and Technology Conferences (Scottsdale, AZ), the 2023 Annual Midwinter Meeting for the Association for Research in Otolaryngology (San Jose, CA), the 2023 Meeting of the Acoustical Society of America (Sydney, AU), and the 2024 International Hearing-Aid Research Conference (Lake Tahoe, CA).

## Learning Outcomes

The reader will understand how the qVCV test was developed and its potential clinical applications for predicting and quantifying greater-than-expected hearing difficulties and hearing aid benefit.

## Appendices (optional)

None

## Supplemental information (optional)

None

